# Association of the LIfestyle for BRAin Health Score and Brain Vascular Damage in a Community-Dwelling Adults Cohort

**DOI:** 10.1101/2023.10.29.23297739

**Authors:** Hang Li, Qi Zhou, Xueli Cai, Yicong Wang, Yingying Yang, Suying Wang, Xia Meng, Jing Jing, Dandan Liu, Yanli Zhang, Yongjun Wang, Yuesong Pan, Yilong Wang

## Abstract

**OBJECTIVE:** To investigate the association of brain vascular damage and the LIfestyle for BRAin Health (LIBRA) dementia risk scores.

**METHODS:** This cross-sectional study was based on the baseline survey from a population-based prospective cohort study. The Polyvascular Evaluation for Cognitive Impairment and Vascular Events (PRECISE) study recruited community-dwelling adults aged 50–75 years based on cluster sampling from Lishui city in China. Eleven modifiable risk or protective factors for dementia collected at baseline survey were combined in the LIBRA score. Eligible participants were scanned by a 3.0T Magnetic resonance imaging scanner. Intracranial atherosclerosis and cerebral small vessel disease (CSVD) burden were assessed.

**RESULTS:** In a total of 3059 individuals, the mean (SD) age was 61.2 (6.7) years; 1630 (53.5%) being women. Compared with the first tertile, subjects with the third tertile of LIBRA score were associated with increased odds of the presence of intracranial atherosclerotic plaque (adj. OR 2.22, 95% CI 1.75 to 2.82, *P* <0.001). A similar association was observed for intracranial atherosclerotic burden (adj. cOR 2.26, 95% CI 1.79 to 2.87, *P* <0.001). Whereas, subjects with the third tertile of LIBRA score were not associated with the presence of CSVD (adj. OR 1.05, 95% CI 0.86 to 1.29, *P* =0.63) or higher CSVD burden (adj. cOR 1.06, 95% CI 0.87 to 1.29, *P* =0.55). Multivariable binary logistic regression analysis for the association between the LIBRA score and the presence of coexistent atherosclerotic plaque and CSVD was also significant adjusting for the nonmodifiable factors age, sex, education, married or cohabiting status, socioeconomic status, and history of stroke (adj. OR 1.70, 95% CI 1.21 to 2.39, *P* = 0.002).

**CONCLUSIONS:** In this study, LIBRA scores were positively associated with the presence and burden of intracranial atherosclerosis, coexistence of atherosclerotic plaque and CSVD. These findings suggested that modifiable dementia score maybe useful in identifying individuals at high risk of brain vascular damage.

## INTRODUCTION

Worldwide, cerebrovascular diseases and dementia are two of the leading contributors to impairment of brain health and neurological disability in older people.^1^ There is an inseparable and intricate connection between dementia and cerebrovascular damage.^2-4^ Most cognitive impairment in the elderly arises from multiple pathologies, of which the vascular component is currently the only treatable and preventable one. ^5,6^ Furthermore, dementia and stroke share similar risk factors, and various lifestyles or risk factors may affect cognitive function through the accumulation of vascular damage.

The LIfestyle for BRAin Health (LIBRA) index is the first validated dementia risk score predominantly supported by modifiable health and lifestyle factors.^7^ The modifiable risk and protective factors for dementia were compiled in a systematic review and Delphi consensus study.^8^ The relative risk of each factor was standardized and weighted to a reference value used to calculate a personalized LIBRA global score. Longitudinal population-based studies suggested that an increase in the global LIBRA score by one point was related to a 13-19% higher risk of developing dementia.^9^ The LIBRA score can be useful as a composite score for stratifying participants into different levels of dementia risk, as well as a surrogate endpoint and surveillance tool to monitor intervention success.^9^ However, it is not yet clear whether the LIBRA scale is related to cerebrovascular damage and its severity.

In this population-based cohort study, we aim to investigate the associations between LIBRA score and brain vascular damage, including atherosclerotic plaque and cerebral small vessel disease (CSVD). In addition, we also aim to observe if the high-risk population with the coexistence of both atherosclerosis and CSVD have higher dementia risk score.

## METHODS

### Study Design and Participants

This study was based on data of the cross-sectional survey of the PolyvasculaR Evaluation for Cognitive Impairment and vaScular Events (PRECISE) study, which is a population-based prospective cohort study with a comprehensive evaluation of atherosclerosis in multiple vascular territories using advanced vascular imaging techniques. The design and baseline data of PRECISE study (NCT03178448) have been previously published.^10^ The protocol received approval from the ethics committee at Beijing Tiantan Hospital and Lishui Hospital. All participants provided written informed consent before enrollment. The baseline data collection of PRECISE was performed from May 2017 to September 2019. Data was analyzed from April to June 2023. This study adhered to the STROBE guidelines (Strengthening the Reporting of Observational Studies in Epidemiology).^11^

### Data Collection and the LIBRA Scale

Data collection for all participants was conducted at Lishui Hospital by trained investigators with standardized questionnaires through face-to-face clinical interviews. The collected data included demographic characteristics (age and gender, education, socioeconomic condition), medical history (hypertension, diabetes, hypercholesterolemia, stroke, coronary artery disease, and kidney disease), and behavior habit (smoking and drinking status, diet, stress, sleep conditions, and physical activity). Additionally, fasting blood samples were obtained to measure the lipid profile, fasting blood glucose (FBG), glycated hemoglobin (HbA1C), and renal function levels.

Individual exposure to modifiable dementia risk factors is calculated using the LIBRA scale, which consists of 12 modifiable risk and protective factors. Each factor is assigned a weight based on its relative risk referring to previous research results.^12^ These weights are then standardized and summed up to yield the final LIBRA score, with higher scores indicating greater dementia risk. In this study, data are available for all LIBRA factors, except for cognitive activity, resulting in a modified LIBRA score with a theoretical range from −2.7 to 12.7. Risk factors include coronary heart disease, diabetes, hypercholesterolemia, hypertension, depression, obesity, smoking, physical inactivity, and chronic kidney disease. Protective factors are low-to-moderate alcohol use and healthy diet. These factors are dichotomized based on previously used cut-offs and Table S1 provides an overview of all individual LIBRA factors, assigned weights, and operationalization in this dataset. Coronary heart disease was based on self-reports of diagnoses made by a physician diagnoses. Hypertension was defined as a SBP > 140 mmHg or DBP > 90 mmHg. Participants were classified as obesity if their body mass index (BMI), calculated as the weight in kilograms divided by height in meters squared, exceeded 30 kg/m^2^. The cut-off point for high blood sugar was glycated hemoglobin (HbA1C) of 6.5% or greater. Hypercholesterolemia was defined as total cholesterol level >5.0 mmol/L or low-density lipoprotein >3.0 mmol/L. The cut-off for renal dysfunction was based on the estimated glomerular filtration rate (eGFR) < 60 ml/min/1.73 m^2^. Protective factor “Low-to-moderate alcohol use” was defined as drinking alcohol occasionally and less than average 20 g/d daily, including non-drinkers. Current smoking was defined as those who smoked at least 1 cigarette per day on average during the last month. A healthy diet was defined as meeting at least 2 of the 3 food recommendations: (1) fruits and vegetables: >4.5 servings (1020 g) per day; (2) fish: ≥two servings (200 g) per week; and (3) sodium: <6 g per day. Physical activity was adjusted according to the International Physical Activity Questionnaire (IPAQ).^13^ Due to the lack of a depression score in the baseline survey of this study, we replaced the depression factor with the results of the survey questionnaire regarding stress. A cut-off point was established based on the sum score ≥2, where each “yes” answer counted as 1 point, out of three questions related to self-reported feeling of stress: moderate to severe life pressure, moderate to severe work pressure, and any stress strike within one year. The modified version of the sum LIBRA score was further categorized into three groups according to the tertiles: The first tertile group represented the low-risk group for dementia, while the third tertile group, with high LIBRA scores, represented the high-risk group for dementia.

### MRI Acquisition and Assessment

Eligible subjects without contraindications for MRI were scanned by a 3.0T scanner (Ingenia 3.0T, Philips, Best, The Netherlands) during the baseline survey. The presence of atherosclerotic plaque was defined as eccentric wall thickening with or without luminal stenosis observed on 3-dimensional time-of-flight magnetic resonance angiography (3-D–TOF MRA), 3-D isotropic high-resolution black-blood T1w vessel wall imaging. The presence of intracranial atherosclerotic stenosis was defined as 50%-99% stenosis or occlusion according to the Warfarin-Aspirin Symptomatic Intracranial Disease Trial [WASID] criteria.^14^ The semi-quantitative atherosclerosis score from 0 to 4 indicates no atherosclerotic plaque, atherosclerotic plaque without evident lumen stenosis or stenosis <50%, stenosis 50%-69%, stenosis 70%-99%, and occlusion.^15^ The intracranial atherosclerotic burdens were calculated by summing the scores of each arterial segment of distal internal carotid, middle cerebral (M1 and M2), anterior cerebral (A1 and A2), posterior cerebral (P1 and P2), basilar and vertebral arteries (V4), which were eventually categorized into scores of 0, 1, 2–3 and ≥4, respectively.^16^

The MRI markers of CSVD, including lacunar infarcts, white matter hyperintensity (WMH), cerebral microbleeds (CMB), and enlarged perivascular spaces (PVS), were defined by the Standards for Reporting Vascular Changes on Neuroimaging Criteria.^17^ The presence and burden of CSVD were evaluated based on a validated CSVD score ranged from 0 to 4, designed by Wardlaw.^18^ Further, we also evaluated modified total CSVD burden using a validated ordinal score that ranged from 0 to 6, described by Rothwell.^19^

Imaging data were collected in DICOM format on discs and centrally adjudicated at Beijing Tiantan Hospital. Two well-trained assessors who were blinded to all clinical data evaluated vascular stenosis and CSVD imaging markers independently and discrepancies were resolved by another senior neurologist. Good interobserver reproducibility was found for the presence of plaque (kappa=0.97), and each CSVD marker between raters (kappa=0.80 for lacune, 0.82 for WMH, 0.90 for PVS and 0.80 for CMB, respectively).

### Statistical Analysis

Continuous variables were presented as the mean with SD or median with IQR, as appropriate, and categorical variable were presented as frequency with percentage. Baseline characteristics among LIBRA score tertile groups were compared using the χ2 test for categorical variables and one-way analysis of variance or the Kruskal–Wallis test for continuous variables. We used multivariable binary logistic regression to examine the relationships between LIBRA score and the presence of intracranial atherosclerotic plaque and CSVD. Initially, the LIBRA scale was treated as a categorical variable, with the first tertile as the reference. Subsequently, it was treated as a continuous variable with increments of 1 standard deviation; For each dependent variable, an unadjusted regression model and two adjusted regression models were conducted. We only adjusted for age and gender in the first modified model. Then, in modified model 2, we further adjusted for years of education, married or cohabiting, socioeconomic status, and history of stroke. For each model, ORs with 95% CIs were calculated. We used multivariable ordinal logistic regression to examine the associations between the LIBRA score and a shift in the direction of a higher intracranial atherosclerotic burden or CSVD burden using the first tertile as the reference. Common ORs with 95% CIs were evaluated. To further assess the pattern and magnitude of the relationship between the LIBRA score and brain vascular damage, full adjusted multivariable binary and ordinal logistic regression models with 4 knots (5th, 35th, 65th, and 95th percentiles) restricted cubic splines for the LIBRA score were performed. Finally, as a sensitivity analysis, we used the modified total CSVD burden, another validated CSVD score method, to reassess the relationship between LIBRA score and intracranial atherosclerotic burden, CSVD, as well as coexistent atherosclerotic plaque and CSVD. A two-sided *P* < .05 was considered to be statistically significant. All analyses were performed using SAS software, version 9.4 (SAS Institute Inc).

## RESULTS

### Baseline Characteristic

The PRECISE study enrolled 3067 community-dwelling adults at baseline. After excluding 6 subjects with missing or poor quality MR images and 2 subjects with missing LIBRA score data, a total of 3059 subjects were included in this analysis (Figure S1). The baseline demographics of the included participants according to tertiles of LIBRA score were shown in Table S2. The mean (SD) age of the participants was 61.2 (6.7) years, and 53.5% (1636) were female. Participants with high LIBRA scores were more likely to be men, with a higher level of BMI, SBP, DBP, FPG, HbA1C, lipid, and creatinine level, more likely to have a medical history of hypertension, diabetes, current smoking, current drinking, and more with physical inactivity, stress, and unhealthy diet.

### Association of LIBRA Score with Intracranial Atherosclerosis and CSVD

Among all the 3059 included subjects, 541 (17.7%) participants had intracranial atherosclerotic plaque, among whom 304 (9.9%), 200 (6.5%), and 37 (1.2%) had intracranial atherosclerotic burden score of 1, 2–3 and ≥4, respectively. There were 933 (30.5%) participants having ≥1 score of CSVD burden (16.6%, 5.6%, 10.2% and 9.8% had presence of confluent WMH, lacunes, CMBs and BG-PVS >10), among whom 678 (22.2%), 176 (5.8%), and 79 (2.6%) had CSVD burden score of 1, 2, and 3–4, respectively. The distribution of intracranial atherosclerotic burden and CSVD burden by tertiles of LIBRA score are presented in Figure 1. From the percentage bar graph, we found that from the first tertile to the third tertile group, with the increase of LIBRA score, the proportion of higher arteriosclerotic plaque burden and higher CSVD burden groups gradually increased.

**Figure 1.**
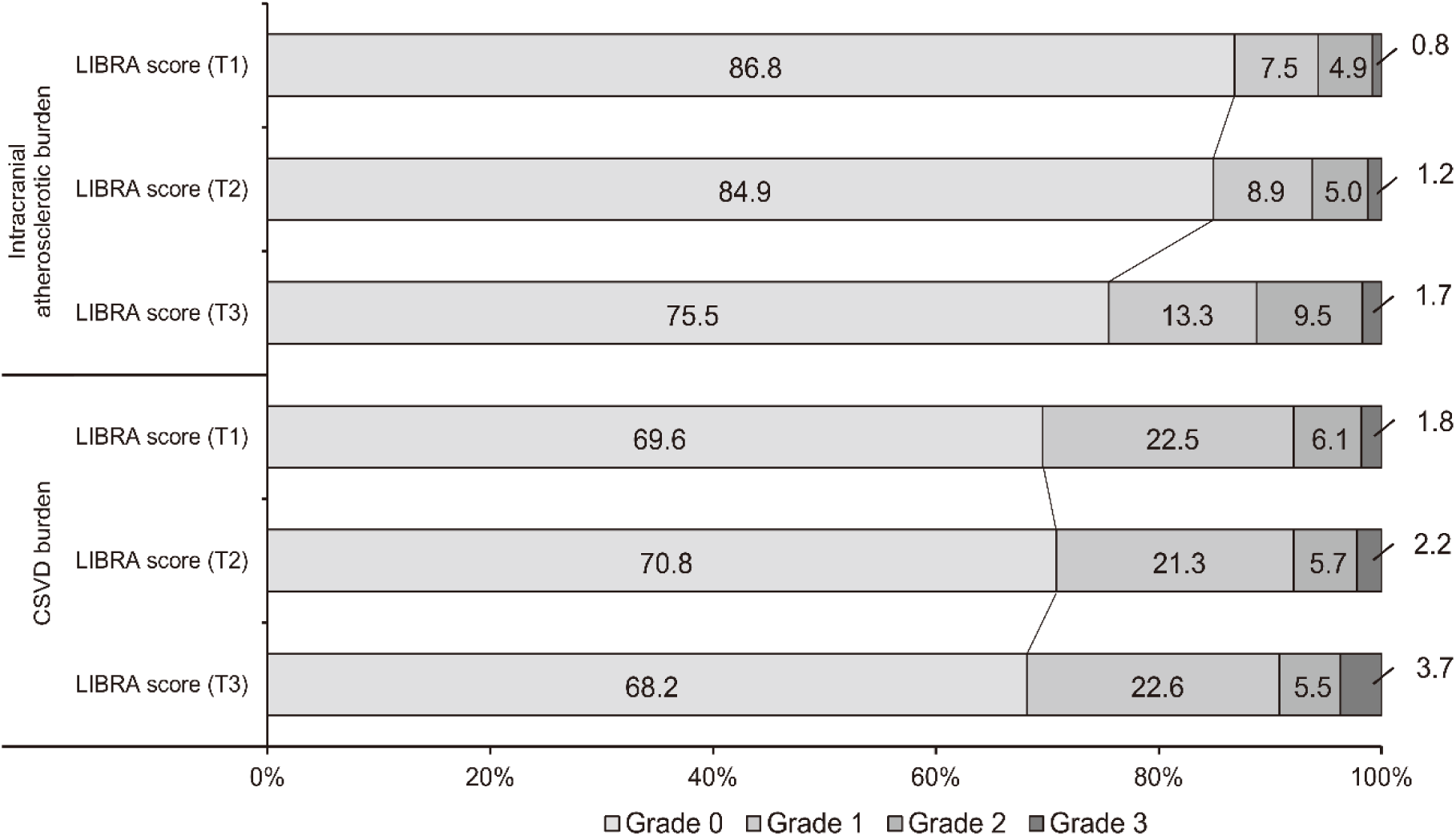
Distribution of Intracranial Atherosclerotic Burden and CSVD Burden by Tertiles of LIBRA. Grades 0–4 indicate intracranial atherosclerotic burden score of 0, 1, 2–3, and ≥4, or total CSVD burden score of 0, 1, 2, 3–4, respectively. CSVD, cerebral small vessel disease; LIBRA, LIfestyle for BRAin health; T1, the lowest LIBRA tertile; T2, the medium LIBRA tertile; T3, the highest LIBRA tertile.

Compared to the first tertile, subjects with the third tertile of LIBRA score were associated with increased odds of the presence of intracranial plaque (adj. OR 2.22, 95% CI 1.75 to 2.82, *P* < 0.001). A similar association was observed for intracranial atherosclerotic burden (adj. cOR 2.26, 95% CI 1.79 to 2.87, *P* < 0.001). Whereas, LIBRA score was not associated with the presence of CSVD (adj. OR 1.05, 95% CI 0.86 to 1.29, *P* = 0.63) or CSVD burden (adj. cOR 1.06, 95% CI 0.87 to 1.29, *P* = 0.55) (Table 1).

**Table 1.** Association of the LIBRA Score and Brain Vascular Damage.

| Brain Vascular Damage | LIBRA score | Unadjusted Model |  | Adjusted Model 1 |  | Adjusted Model 2 |  |
| --- | --- | --- | --- | --- | --- | --- | --- |
|  |  | OR/cOR(95% CI) | *P* | OR/cOR(95% CI) | *P* | OR/cOR(95% CI) | *P* |
| Presence of intracranial plaque | T1 | Ref |  | Ref |  | Ref |  |
|  | T2 | 1.17 (0.91-1.51) | 0.21 | 1.21 (0.94-1.56) | 0.14 | 1.19 (0.92-1.54) | 0.19 |
|  | T3 | 2.13 (1.70-2.68) | <0.001 | 2.28 (1.81-2.89) | <0.001 | 2.22 (1.75-2.82) | <0.001 |
|  | per SD | 1.40 (1.27-1.53) | <0.001 | 1.44 (1.30-1.58) | <0.001 | 1.43 (1.29-1.57) | <0.001 |
|  | P trend |  | <0.001 |  | <0.001 |  | <0.001 |
| Intracranial atherosclerotic burden | T1 | Ref |  | Ref |  | Ref |  |
|  | T2 | 1.17 (0.91-1.50) | 0.22 | 1.21 (0.94-1.57) | 0.14 | 1.20 (0.93-1.55) | 0.16 |
|  | T3 | 2.13 (1.70-2.67) | <0.001 | 2.32 (1.84-2.93) | <0.001 | 2.26 (1.79-2.87) | <0.001 |
|  | per SD | 1.40 (1.28-1.54) | <0.001 | 1.45 (1.32-1.60) | <0.001 | 1.44 (1.31-1.59) | <0.001 |
|  | P trend |  | <0.001 |  | <0.001 |  | <0.001 |
| Presence of CSVD | T1 | Ref |  | Ref |  | Ref |  |
|  | T2 | 0.95 (0.78-1.15) | 0.58 | 0.94 (0.76-1.15) | 0.54 | 0.93 (0.76-1.14) | 0.50 |
|  | T3 | 1.07 (0.89-1.29) | 0.48 | 1.06 (0.87-1.30) | 0.55 | 1.05 (0.86-1.29) | 0.63 |
|  | per SD | 1.05 (0.97-1.13) | 0.21 | 1.05 (0.96-1.14) | 0.27 | 1.05 (0.96-1.14) | 0.30 |
|  | P trend |  | 0.48 |  | 0.54 |  | 0.60 |
| CSVD burden | T1 | Ref |  | Ref |  | Ref |  |
|  | T2 | 0.95 (0.79-1.15) | 0.63 | 0.93 (0.76-1.14) | 0.48 | 0.92 (0.75-1.13) | 0.42 |
|  | T3 | 1.09 (0.91-1.30) | 0.37 | 1.09 (0.89-1.32) | 0.40 | 1.06 (0.87-1.29) | 0.55 |
|  | per SD | 1.06 (0.98-1.14) | 0.13 | 1.06 (0.98-1.15) | 0.13 | 1.06 (0.98-1.15) | 0.17 |
|  | P trend |  | 0.37 |  | 0.40 |  | 0.53 |
| Coexistent plaque + CSVD | T1 | Ref |  | Ref |  | Ref |  |
|  | T2 | 1.10 (0.77-1.57) | 0.60 | 1.12 (0.78-1.61) | 0.55 | 1.12 (0.78-1.63) | 0.53 |
|  | T3 | 1.69 (1.23-2.33) | 0.001 | 1.79 (1.28-2.49) | <0.001 | 1.70 (1.21-2.39) | 0.002 |
|  | per SD | 1.32 (1.16-1.50) | <0.001 | 1.35 (1.18-1.54) | <0.001 | 1.32 (1.15-1.52) | <0.001 |
|  | P trend |  | <0.001 |  | <0.001 |  | 0.002 |

Note: LIBRA risk groups: T1:<0.4, T2: 0.4 to 2.1, T3: ≥ 2.1; LIBRA score theoretical range: -2.7 to 12.7; observed range: -2.7 to 8.5.
Unadjusted Model: crude model. Adjusted Model 1: adjusted for age, and gender. Adjusted Model 2: adjusted for model1+ years of education, married or cohabiting, socioeconomic status, and history of stroke.
OR and 95% CI for the presence of intracranial plaque, and CSVD by binary logistic regression, whereas cOR and 95% CI for intracranial atherosclerotic burden and CSVD burden by ordinal logistic regression. The degree of intracranial atherosclerotic burden was graded as scores of 0, 1, 2-3, and ≥ 4, respectively. The degree of CSVD burden (Wardlaw) was graded as scores of 0, 1, 2, and 3-4, respectively. One point was allocated for the presence of lacunes, WMH burden (PV-WMH Fazekas 3 or deep-WMH Fazekas 2-3), presence of CMBs, and moderate-to-severe basal ganglia PVS (BG-PVS) (N>10).
CSVD, cerebral small vessel disease; WMH, white matter hyperintensity; PV-WMH, paraventricular-white matter hyperintensity; BG-PVS, basal ganglia-perivascular spaces; OR, odds ratio; cOR, common OR; Ref., reference; LIBRA, Lifestyle for BRAin health; T1, the lowest LIBRA tertile; T2, the medium LIBRA tertile; T3, the highest LIBRA tertile.

The third tertile of LIBRA score was associated with increased odds of the presence of confluent WMH (Fazekas score: d-WMH 2-3 or PV-WMH 3) (T3 vs T1: adj. OR 1.48, 95% CI 1.16 to 1.90, P < 0.01). However, it was not significant for lacunes ≥1 (T3 vs T1: adj. OR 1.22, 95% CI 0.82 to 1.82, P = 0.33), cerebral microbleed ≥1 (T3 vs T1: adj. OR 0.81, 95% CI 0.60 to 1.08, P = 0.15), or BG-PVS >10 (T3 vs T1: adj. OR 0.95, 95% CI 0.70 to 1.30, P = 0.75) (Figure 2).

**Figure 2.**
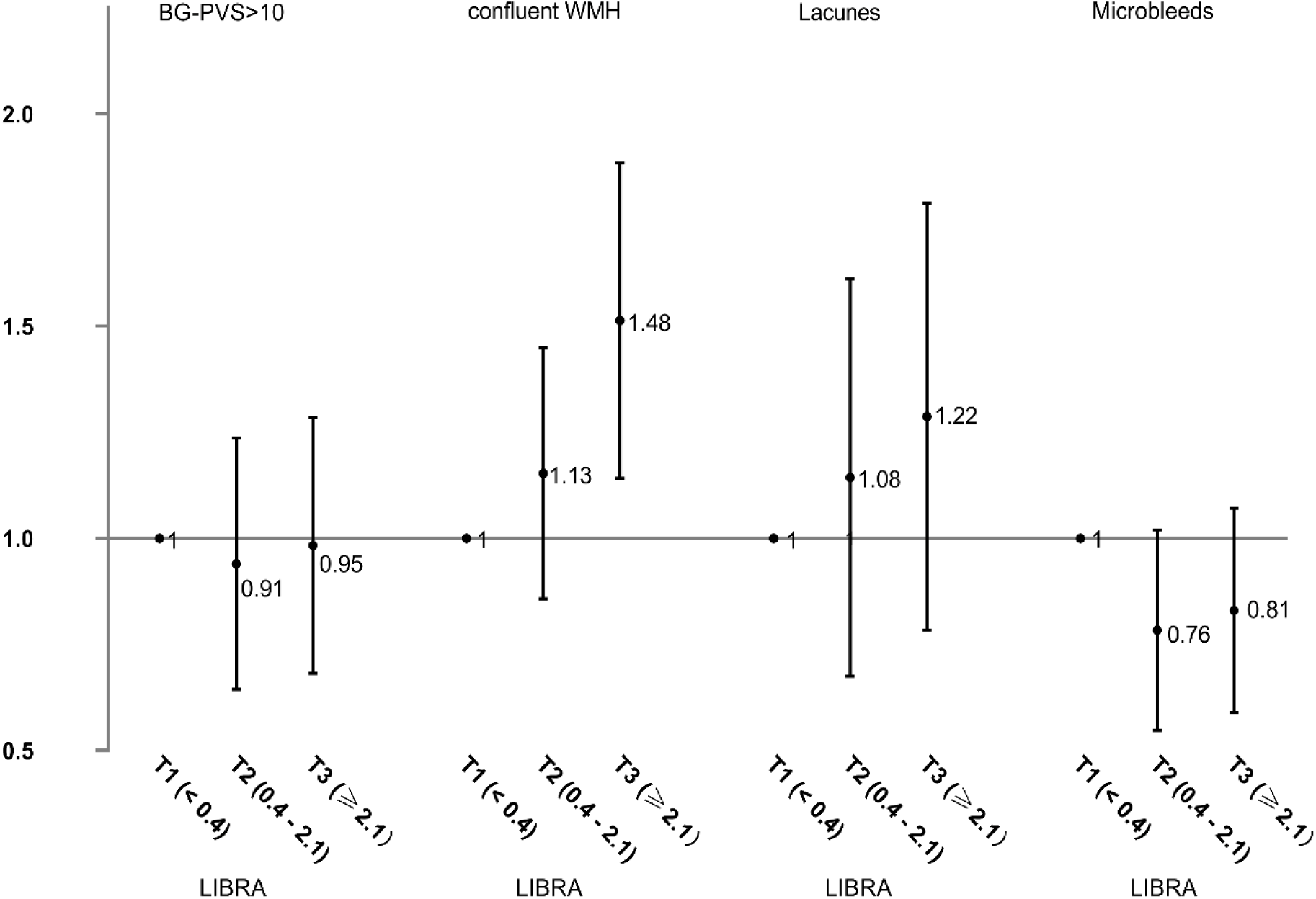
Forest Plots for the Association of the LIBRA Score and CSVD Imaging Markers. The plots showed the OR and their 95% CIs, adjusted for age, gender, years of education, married or cohabiting, socioeconomic status, and history of stroke. CSVD, cerebral small vessel disease; BG-PVS, basal ganglia-perivascular spaces; WMH, white matter hyperintensity; OR, odds ratio; LIBRA, LIfestyle for BRAin health; T1, the lowest LIBRA tertile; T2, the medium LIBRA tertile; T3, the highest LIBRA tertile.

### Association of LIBRA Score with Coexistent Intracranial Atherosclerosis and CSVD

Distribution of coexistent intracranial atherosclerosis and CSVD by tertiles of LIBRA score were presented in Figure 3. Compared with the first tertile group, the third tertile group with a high LIBRA score had the highest proportion of coexistent atherosclerotic plaque and CSVD, followed by atherosclerotic plaque alone and the CSVD alone, and the lowest proportion of the population with neither arteriosclerosis nor CSVD imaging markers.

**Figure 3.**
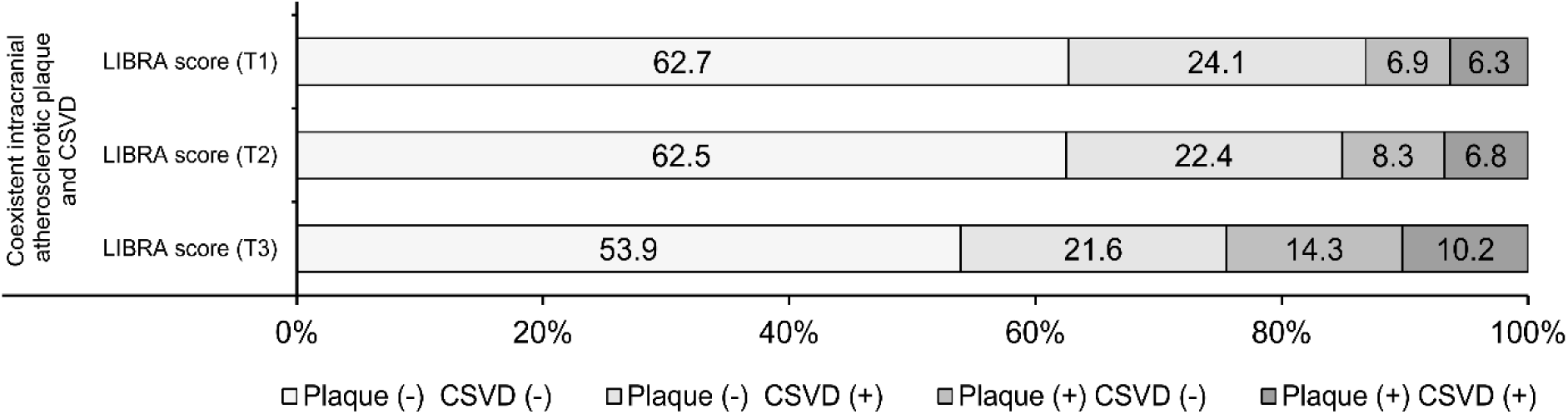
Distribution of Coexistent Intracranial Atherosclerotic Plaque and CSVD by Tertiles of LIBRA. CSVD, cerebral small vessel disease; LIBRA, LIfestyle for BRAin health; T1, the lowest LIBRA tertile; T2, the medium LIBRA tertile; T3, the highest LIBRA tertile.

Multivariable binary logistic regression was analyzed for the association between the LIBRA score and coexistent atherosclerotic plaque and CSVD (T3 vs T1: adj. OR 1.70, 95% CI 1.21 to 2.39, *P* = 0.002) after adjustment for the nonmodifiable factors age, sex, education, married or cohabiting status, socioeconomic status, and history of stroke (Table 1). Using a binary or ordinal logistic regression with a restricted cubic spline for LIBRA score, we found that subjects with higher LIBRA scores had higher odds of the presence of intracranial atherosclerotic plaque and higher intracranial atherosclerotic burden, but not associated with CSVD burden. Population with higher LIBRA scores had higher odds of the presence of coexistent atherosclerotic plaque and CSVD (Figure 4).

**Figure 4.**
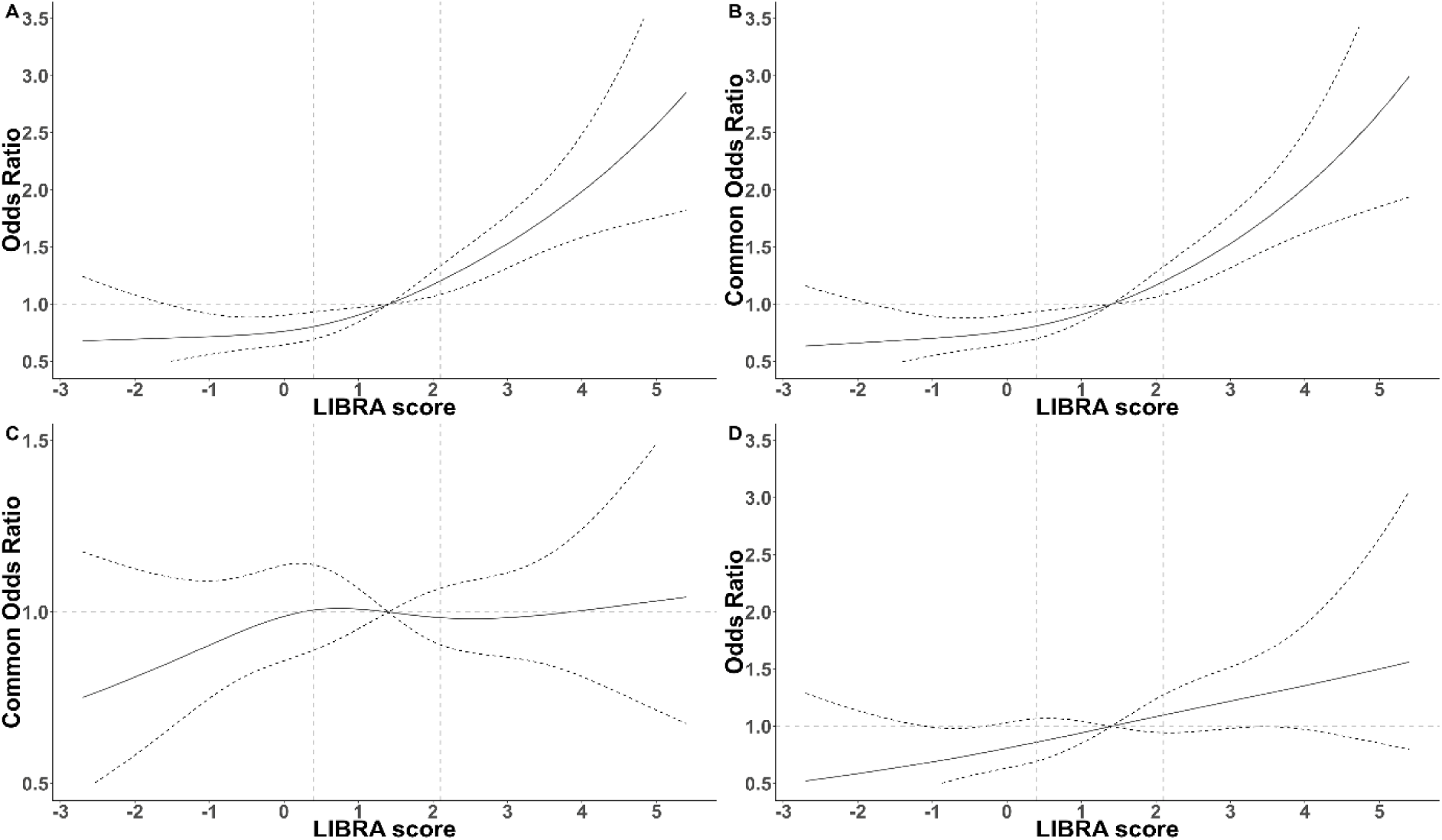
Association of LIBRA Score and Brain Vascular Damage. Multivariable-adjusted ORs for the presence of intracranial plaque(A) and coexistent intracranial plaque + CSVD (D), based on restricted cubic spines with 4 knots (5th, 35th, 65th, and 95th percentiles) of LIBRA score. Multivariable-adjusted common ORs for the intracranial atherosclerotic burden (B) and CSVD burden (C), based on restricted cubic spines with 4 knots (5th, 35th, 65th, and 95th percentiles) of LIBRA score. All models adjusted for age, gender, years of education, married or cohabiting, socioeconomic status, and history of stroke. CSVD, cerebral small vessel disease; LIBRA, LIfestyle for BRAin health score; OR, odds ratio.

Finally, as a sensitivity analysis, we used the modified CSVD burden score ranging from to 6 to reassess the relationship between LIBRA score and CSVD, as well as coexistent atherosclerosis and CSVD. After adjusting for the same factors as above, we reached the similar conclusion. (Table S3).

## DISCUSSION

In a large-scale community-based population, we discovered that brain vascular damages such as atherosclerotic plaque, coexistence of both atherosclerotic plaque and CSVD were associated with a higher LIBRA score. The LIBRA score was originally developed to estimate an individual’s risk for developing dementia in cognitively intact adults; however, our results indicated the LIBRA score might also be useful in predicting vascular damage and its severity.

Previous research has shown that a higher cardiovascular risk burden was associated with an increased risk of dementia. Cerebral infarctions and atherosclerosis might underlie this association. In this study, we found that an elevated LIBRA score was positively associated with the presence and burden of intracranial atherosclerosis. According to a meta-analysis study, consistent with those in the LIBRA score, the risk factors for arteriosclerosis included advanced age, metabolic syndrome, diabetes mellitus, hypertension, and dyslipidemia.^20^ Increased attention has been focused on a constellation of novel lifestyle factors, including diet quality, physical activity, cigarette smoking, alcohol consumption, and body weight.^21,22^ Atherosclerosis was an age-related condition that constituted a major risk factor for cerebral microangiopathies. Vascular damage caused by atherosclerosis can lead to microadenomas, microaneurysms, and even stenosis or obstruction of the vessel,^23,24^ while stenosis and hypoperfusion caused incomplete ischemia lesions evidenced by neuroimaging as white matter hyperintensity.

Previous research has shown that the higher Life’s Simple 7 (LS7) score was associated with the lower presence of CSVD and a lower total CSVD score.^25^ However, in this study, the relationship between LIBRA score and CSVD was not significant. There were some reasons to explain these differences: First, although both the LS7 and LIBRA scores contained similar modifiable risk factors, the objectives, and scoring methods were different. Second, the LIBRA score included several additional factors including kidney disease, heart disease, and alcohol consumption, which may be more associated with large arterial atherosclerosis than small vessel disease. The mechanism of cognitive impairment caused by CSVD, besides these modifiable risk factors, includes vascular endothelial dysfunction, chronic vascular inflammation, cerebrovascular reactivity abnormalities,^26^ blood–brain barrier damage,^27^ and meningeal lymphatic system dysfunction. The LIBRA score, only considering the impact of modifiable factors on dementia, may underestimate the risk of dementia in the CSVD population.

Aligned with previous studies,^28^ participants with higher LIBRA scores denoting higher dementia risk, had higher volumes of WMH. WMH load was associated with Alzheimer disease risk factors even in cognitively unimpaired subjects.^29^ WMH progression was associated with the development of probable dementia or mild cognitive impairment.^30^ All these suggested that WMH volume may partially mediate the association between LIBRA score and cognition. The relationship between WMH and dementia risk factor seemed to be stronger than other CSVD imaging markers or CSVD total burden.

Several vascular risk factors, such as hypertension, obesity, and diabetes had been shown to exacerbate neurovascular impairments and thus increase vascular dementia prevalence.^23^ Population with higher LIBRA scores had higher odds of the presence of coexistent atherosclerosis and CSVD, suggesting that the population with more modifiable risk factors may have more serious accumulated vascular damage and a higher risk of dementia. Considering that approximately 90% of stroke is attributable to modifiable risk factors,^31^ preventing dementia by controlling vascular risk factors and cerebrovascular diseases may be promising.^32^

It is important to acknowledge the limitations of this study. First, this study was a cross-sectional analysis of a baseline survey of the PRECISE cohort. Potential unmeasured confounders and reverse causation may exist in an observational study. Second, the absence of the LIBRA factor cognitive activity, the strongest protective factor, could weaken the predictive value of the LIBRA score. However, the purpose of this study was to investigate the correlation between modifiable risk factors and vascular damage, so this modified LIBRA score may be more meaningful for this study. Third, while data on most LIBRA factors were available and intact in this dataset, some variables could only be included via proxy measures, for example, healthy diet. Fourth, this study did not specifically investigate the contribution of each LIBRA component item to vascular damage, considering that any component indicator cannot represent the overall risk of dementia. Finally, since participants in this cohort were recruited from Lishui City and restricted to elderly adults in China, further large-scale investigation is required before extrapolation of the findings to other populations.

## CONCLUSIONS

With comprehensive evaluation of vascular damage using advanced vascular imaging techniques, we found that a higher LIBRA score was positively associated with intracranial atherosclerotic plaque, and the presence of coexistent atherosclerotic plaque and CSVD. In addition to dementia, the LIBRA score might also be useful in predicting brain vascular damage and its severity.

## Data Availability

The data supporting the findings of this study are available from the corresponding author upon reasonable request.

## Acknowledgment

We are grateful for the assistance of the staff, the participants, and the other investigators of the PRECISE study.

## Sources of Funding

This study was supported by the National Natural Science Foundation of China (No.81825007), Beijing Outstanding Young Scientist Program (No. BJJWZYJH01201910025030), Capital’s Funds for Health Improvement and Research (2022-2-2045), Youth Beijing Scholar Program (No.010), Key Science & Technologies R&D Program of Lishui City (2019ZDYF18), Dalian Medical Science Research Program (211017).

## Ethics Approval and Consent to Participate

According to the principles mentioned in the Declaration of Helsinki, the ethics committees of Beijing Tiantan Hospital (IRB Approval Number: KY2017010-01) and Lishui hospital (IRB Approval Number: 2016-42) approved the study protocol. Written informed consent was obtained from all participants in this study.

## Disclosures

None.

## Supplemental Material

Tables S1–S3

Figure S1

## Notes

### Competing Interest Statement

The authors have declared no competing interest.

### Clinical Trial

ClinicalTrials.gov Registry (NCT03178448)

### Clinical Protocols

https://doi.org/10.1136/svn-2020-000411

https://svn.bmj.com/content/svnbmj/6/1/e000411.full.pdf

### Author Declarations

The protocol of the PRECISE study was approved by ethicscommittee at Beijing Tiantan Hospital (IRB approval number: KY2017-010-01) and ethics committee at Lishui Hospital (IRB approval number: 2016-42). All participantsprovided written informed consents before entering the study.

